# Acceleration of regional, racial and gender disparities in drug overdoses across the United States driven by COVID-19 and fentanyl use

**DOI:** 10.1101/2022.06.22.22276766

**Authors:** Maria R. D’Orsogna, Lucas Böttcher, Tom Chou

## Abstract

**Objectives:** To examine trends in drug overdose deaths by gender, race and geography in the United States during the period 2013-2020.

**Methods:** We used the final National Vital Statistics System multiple cause-of-death mortality files to extract crude rates by gender and race and to calculate the male-to-female ratios of crude rates between 2013 and 2020. We established 2013-2019 temporal trends for four major drug types: psychostimulants with addiction potential (T43.6, such as methamphetamines); heroin (T40.1); natural and semi-synthetic opioids (T40.2, such as those contained in prescription pain-killers); synthetic opioids (T40.4, such as fentanyl and its derivatives) through a quadratic regression and determined whether changes in the pandemic year 2020 were statistically significant. We also identified states, race and gender categories most impacted by drug overdoses.

**Results:** Nationwide, the year 2020 saw statistically significant increases in overdoses for all drug categories except heroin, surpassing predictions based on 2013-2019 trends. Crude rates for Blacks of both genders surpassed those for Whites for fentanyl and psychostimulants in 2018 creating a gap that widened through 2020. In some regions mortality among Whites decreased while overdose rates for Blacks kept rising. The largest 2020 mortality statistic is for Black males in the District of Columbia, with a record 134 overdoses per 100,000 due to fentanyl, 9.4 times more than the fatality rate among White males. Male overdose crude rates in 2020 remain larger than those of females for all drug categories except in Idaho, Utah and Arkansas where crude rates of overdoses by natural and semisynthetic opioids for females exceeded those of males.

**Public Health Implications:** Drug prevention, mitigation and no-harm strategies should include racial, geographical and gender-specific efforts, to better identify and serve at risk groups.

---

According to the Centers for Disease Control and Prevention (CDC) and the National Vital Statistics System (NVSS) 91,798 people died from injury or poisoning from drugs of abuse (opioids and psychostimulants) in 2020, corresponding to a 32% rise over 2019. Provisional mortality rates for 2021 point to further rises in fatal overdoses [1]. The media has greatly discussed these increases, which are bolstered by other reports of rising non-fatal drug overdoses, or requests for drug-related emergency medical services starting in 2020 [2].

The recent overdose surge has been attributed to the many mental health stressors and uncertainties associated with COVID-19, and to pandemic-related disruptions to the drug supply that resulted in higher availability of lethal, illicit synthetic opioids such as fentanyl [3]. However, drug abuse and age-adjusted overdose mortality rates have been steadily increasing over the past 20 years. In particular, the year 2013 is considered the beginning of a “third wave” of overdose fatalities [4,5] marked by a shift from deaths due to prescription opioids towards deaths from synthetic opioids, primarily illicit fentanyl. Recent years have also seen more efforts to better understand differences among socio-economic, racial and gender groups in how, when and why drugs are first sought, how they are consumed, and what no-harm options might be the most effective [6--8]. However, few studies have provided a combined, in-depth analysis of gender, geographical and racial differences in drug-induced deaths, even prior to the pandemic [9]. For example, recent reports show that in 2020 the nationwide drug-induced crude rate for Blacks surpassed that of Whites for the first time since 1999 [10]. While interesting for its many societal implications, this analysis does not differentiate by drug type, gender or geography, hindering localized and/or specific intervention efforts. We aim to fill this gap by investigating mortality patterns during the third wave of drug overdoses in the United States, from 2013 to 2020. We focus on four major classes of drugs and verify whether the values recorded for the pandemic year 2020 are statistically significant compared to years prior.

## METHODS

Overdose death rates for all years between 2013 and 2020 were obtained from the CDC WONDER database which tallies mortality, cause of death, and other relevant demographic data in the United States [11]. Overdoses were identified as drug poisoning as listed by the International Classification of Diseases, 10th Revision (ICD-10) using codes X40-X44 (unintentional), X60-X64 (suicide), X85 (homicide), or Y10-Y14 (undetermined intent). We do not distinguish among these categories and include all above manners of death in our evaluations. The type of drugs involved in these overdoses are listed via additional cause of death codes. We focus on the T40 class (poisoning by narcotics and psychodysleptics) which includes the relevant subclasses T40.1 (heroin), T40.2 (natural and semisynthetic opioids such as morphine and oxycodone, typically found in prescription pain-killers) and T40.4 (synthetic opioids other than methadone, such as fentanyl). Another relevant subclass is overdoses by psychostimulants with abuse potential (T43.6), a category which is listed under class T43 (poisoning by psychotropic drugs). Included in T43.6 are methamphetamines, amphetamines and 3,4-methylenedioxymethamphetamine (MDMA, ecstasy). For simplicity we will refer to the four categories as heroin (T40.1), prescription opioids (T40.2), fentanyl (T40.4) and methamphetamines (T43.6). Since all overdoses in WONDER are fatal, we refer to overdose deaths as simply overdoses. We do not consider overdoses from any other substance such as antidepressants, nor poisoning by alcohol. Crude rates for a specific demographic category are defined as the number of fatalities of that category divided by the mid-year population of the same category, multiplied by the typical factor of 100,000. Since WONDER lists male and female genders only, we use a binary gender classification. We further stratify our data by the four Census Regions listed in WONDER: the Northeast (Region 1), the Midwest (Region 2), the South (Region 3) and the West (Region 4). Since data are also catalogued by state plus the District of Columbia, we examine all 51 jurisdictions and refer to them collectively as “states”. The WONDER database also lists four racial categories (White, Black or African American, Asian or Pacific Islander, American Indian or Alaska Native) but only two (White and Black or African American) experienced enough fatalities to achieve statistical significance, so we focus mostly on them. For each geographical or racial category we consider the crude rates for females and males and derive the male-to-female crude ratio by dividing the two. We identify 2013-2019 trends and determine whether 2020 data are statistical outliers. To do this, we first fit the 2013-2019 data to a quadratic form, which allows for possible non-monotonic behaviors. We then calculate the standard deviation of the residuals between the fitted trends and the data for 2013-2019. If the residual corresponding to 2020 is three times beyond the value of the standard deviation, the 2020 value is considered an outlier.

## RESULTS

Nationwide, fentanyl (T40.4) and methamphetamines (T43.6) overdose crude rates increase monotonically for both genders over the 2013-2019 timeframe as shown in Figure 1. For both drug classes and genders, the 2020 values are statistical outliers, as they are in excess of three standard deviations of the residuals above the quadratic regression fit. In particular, for T40.4 male overdose rates in 2020 exceed projections by 30%; female rates by 28%. For T43.6 male rates exceed projections by 19%; female rates by 18%. The T40.4 male-to-female overdose ratio increases in an approximate linear manner from 2013 to 2016, settling around 2.6 and slightly increasing thereafter. At 2.8, the 2020 value is consistent with this trend. For T43.6 the male-to-female ratio is relatively unform over the 2013-2020 period and fluctuates between 2.3 and 2.6. This suggests that while 2020 saw a dramatic nationwide increase in the number of deaths due to fentanyl and methamphetamines, the impact was similar across genders.

**Figure 1:**
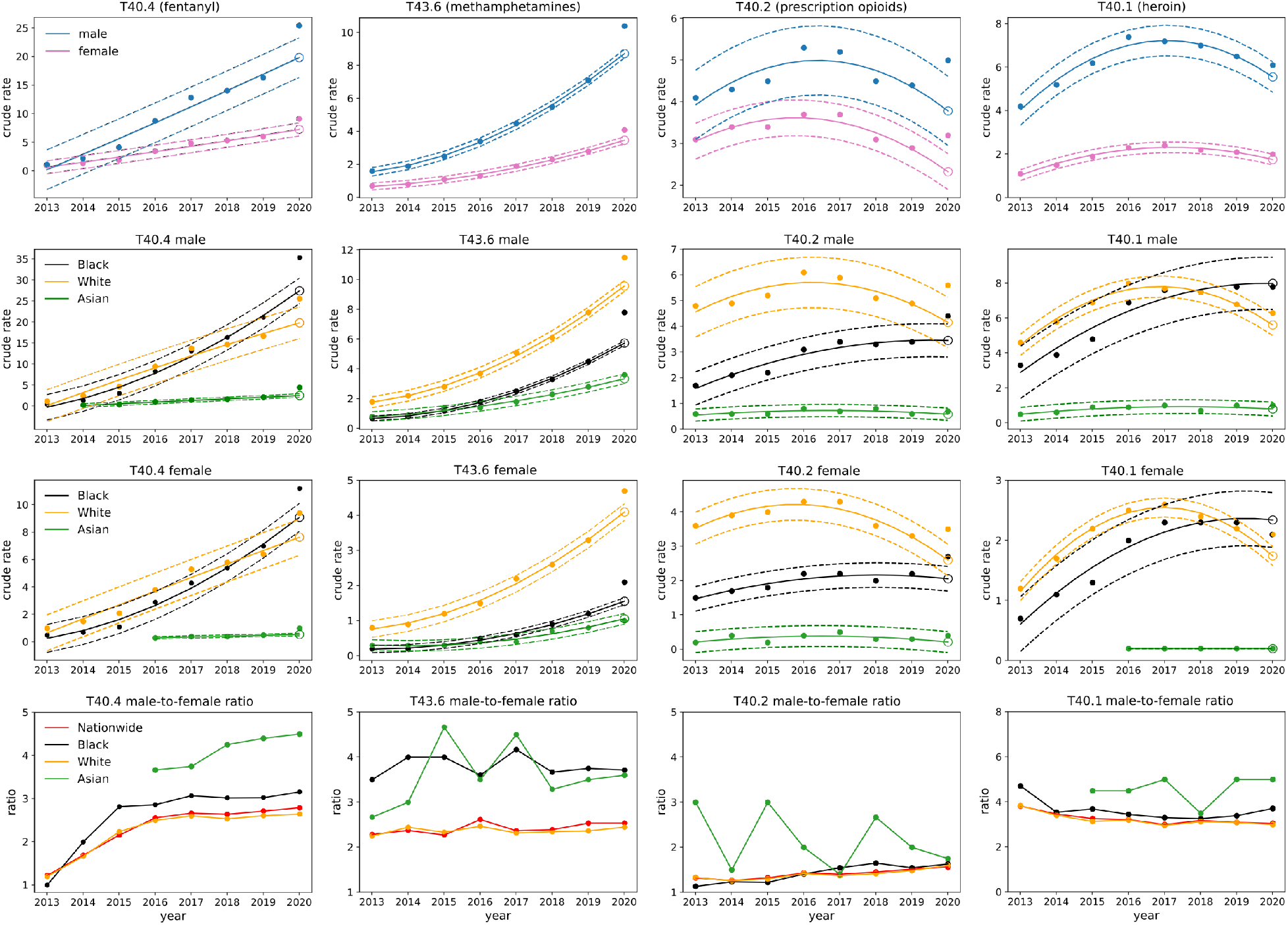
Nationwide gender and race stratified crude rates for synthetic opioids (T40.4, fentanyl), psychostimulants with abuse potential (T43.6, methamphetamines), semi-synthetic opioids (T40.2, prescription opioids) and heroin (T40.1), for the year 2020. Solid lines in the upper three rows represent the 2013-2019 quadratic regression; dotted lines correspond to values three times above and below the regression; the open circle is the 2020 projection. Top row: results are stratified by gender. Note the increasing 2013-2019 trend and the 2020 value exceeding projections for T40.4 and T43.6 for both genders; crude rates for T40.2 and T40.1 have been decreasing but rose again in 2020, with significant rises especially in T40.2 for both genders. Second (third) row from top: results are stratified by race for males (females). For fentanyl and heroin the Black male crude rate surpassed that of Whites in 2018. Last row: Male-to-female ratio for all races. The ratio for Blacks is higher than the ratio for Whites in all categories except for T40.2. We include results for Asians in years when data is available; although crude rates are the lowest in the nation, this is the race with the largest male-to-female ratio in all drug categories.

Further stratifying the data reveals racial disparities for T40.4 that can be seen in Figure 1. Male crude rates for Blacks and Whites were comparable until 2018 (even slightly higher for Whites); the Black male overdose rate exceeded that of White males for the first time in 2019 and in 2020 rose much higher, by about 38%. Similar trends are observed for females: the crude rate for Whites exceeded the rate for Blacks until 2019, but in 2020 the crude rate for Black females surpassed that of White females by 19%. Racial disparities are also observed among the male-to-female ratio of crude rates. All ratios grow until 2015 and later stabilize, however, the Black male-to-female ratio increased more than that of Whites reaching 3.2 in 2020, higher than the 2.6 ratio of Whites. Additional geographical stratifications (not shown) reveal that the Northeast and the Midwest are the most affected by fentanyl mortality for all genders and races, primarily Black males.

The T43.6 overdose rate is higher among Whites than Blacks throughout 2013-2020. However the 2020 male-to-female ratio among Blacks is 3.8, much higher than the male-to-female ratio for Whites, roughly 2.4. A geographic analysis reveals that the highest T43.6 crude rates for all races and genders are in the West.

Nationwide trends for prescription opioids (T40.2) and heroin (T40.1) overdoses display non-monotonic trends: mortality increased between 2013 and 2016 for both genders, and began decreasing in 2017. However, while the 2020 rates for T40.1 remain consistent with downward predictions, those for T40.2 show a statistically significant upward shift, with males surpassing them by 32% and females by 37%. The male-to-female overdose ratio is relatively stable at around 3.1 for T40.1, and is slowly increasing for T40.2 reaching 1.6 in 2020, the lowest of all drug categories.

Stratification by race shows that nationwide Black mortality rates did not decline for either T40.2 or T40.1 over the 2013-2019 interval but instead increased for both genders. In particular, for heroin the nationwide Black male overdose rate was less than that of White males until 2017. After the two reached similar levels in 2018, the crude rate among Blacks kept increasing at a higher rate than among Whites, until in 2020 the crude rate for Blacks exceeded that of Whites by 24%. The Northeast and the Midwest exhibited the highest heroin overdose rates; in both regions male Black mortality has increased or remained stable since 2013, similarly for Black females in the Northeast. Conversely, heroin overdose rates for Whites in these regions declined, especially in the Midwest (not shown). For T40.2 although nationwide overdose rates are larger for Whites than Blacks, in the Northeast White male and female mortalities for both races are decreasing, but are increasing dramatically for Black males (not shown). The male-to-female ratio for each race is slightly larger for Blacks than Whites for both T40.1 and T40.2.

Taken together, these results suggest that the most significant 2020 nationwide changes are the statistically significant increases in mortality due to fentanyl and methamphetamines and a statistically significant 2020 resurgence in prescription opioid deaths for both genders. Despite a non-statistically significant increase in 2020, heroin overdose rates have been declining nationwide since 2017, but not among Blacks. Nationwide overdose rates for Blacks have surpassed that of Whites for fentanyl for both genders, and for heroin for males.

Stratification by gender, race and state does not allow for a consistent analysis over the 2013-2019 range due to insufficient data for many jurisdictions and drug classes so we focus on comparing 2019 data to 2020 data.

In Figure 2 we show geographical variations in the male and female crude rates in 2019 and 2020 within the United States. Large increases for T40.4 and T43.6 crude rates are observed in almost all states, consistent with national trends. The picture is more varied for T40.1 and T40.2 for which a decrease in overdoses is observed across several states. Some display large gender disparities, such as Georgia, where heroin crude rates for females increased by more than 100% but only slightly for males between 2019 and 2020. Similar gender patterns are observed for T40.2 in Minnesota. T40.2 crude rates are instead much larger for males than females in West Virginia and Louisiana.

**Figure 2:**
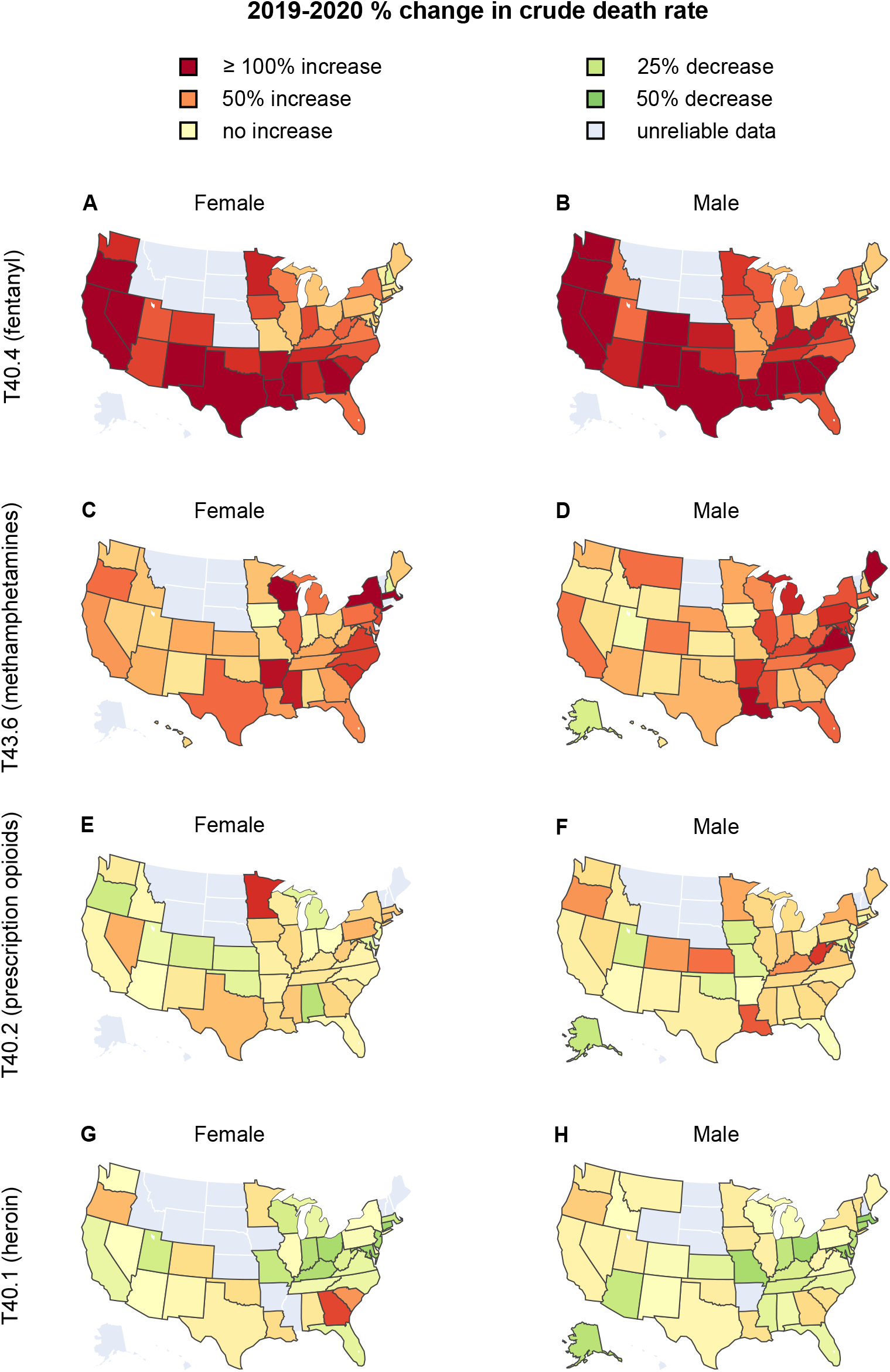
Percentages of change to the crude rate between 2019 and 2020 stratified by gender for fentanyl (T40.4, panels A,B), methamphetamines (T43.6, panels C,D), prescription opioids (T40.2, panels E,F), and heroin (T40.1, panels G,H), in the United States. In states shown in dark red, crude rates more than doubled while green indicates that crude rates decreased between 2019 and 2020.

Gender-stratified crude rates are shown for all states in Figure 3. For all drug classes examined except heroin, the largest crude rates are in West Virginia and Delaware. For heroin, the largest female crude rates are found in Delaware, New Jersey, and West Virginia; the largest male ones in the District of Columbia, Delaware, and New Jersey.

**Figure 3:**
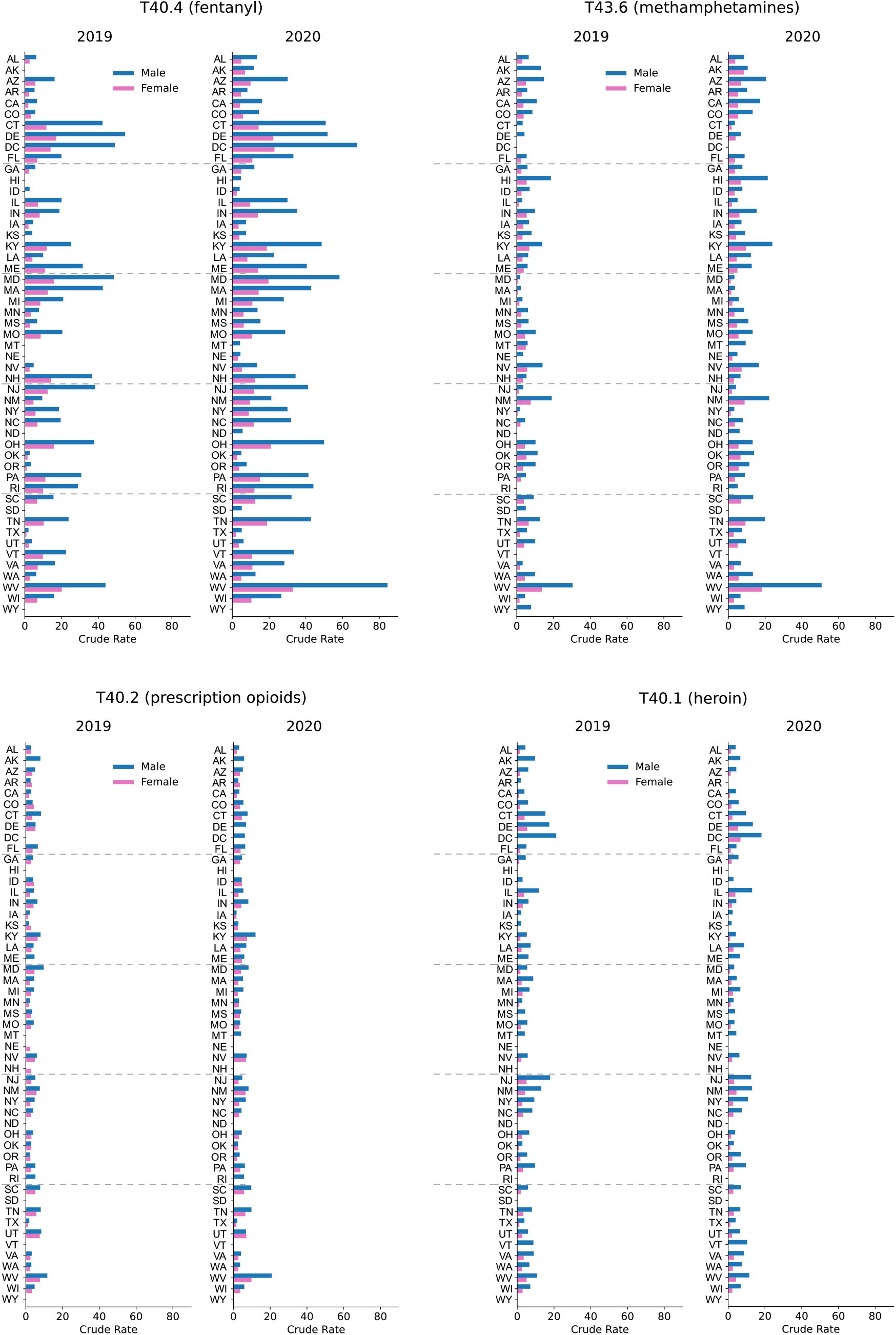
Gender-stratified, crude rates across the United States associated with fentanyl (T40.4), methamphetamines (T43.6), prescription opioids (T40.2), heroin (T40.1) for the years 2019 and 2020. Note the gender imbalances and increase in mortality for West Virginia from 2019 to 2020 for all drug categories except heroin.

As observed above, if only gendered overdoses are considered, the largest 2020 fentanyl crude rates are in West Virginia (84 male and 33 female). Upon differentiating by race, dramatic disparities emerge, primarily in the District of Columbia, as can be seen in Figure 4. Here, until 2015 there was an insufficient number of T40.4 overdoses for either gender or race, however in 2016 a large spike of Black male deaths emerged (57 cases per 100,000) leading to a record crude rate of 134 in 2020 (+35% with respect to 2019) for males and 43 (+56% with respect to 2019) for females. Both are the highest in the nation in 2020 and 2019. The only significant T40.4 datapoint in the District of Columbia for Whites over the entire 2013-2020 period is a male crude rate of 14 in 2020. This implies that the vast majority of fentanyl victims in the District of Columbia are Black males, and that in 2020 there were 9.4 times more fatalities among Blacks than among Whites. Ever since fentanyl deaths were first tallied in the District of Columbia in 2016, Black male crude rates have been larger than those of Whites (and Blacks) in West Virginia, the state with the largest overall male fentanyl mortality.

**Figure 4:**
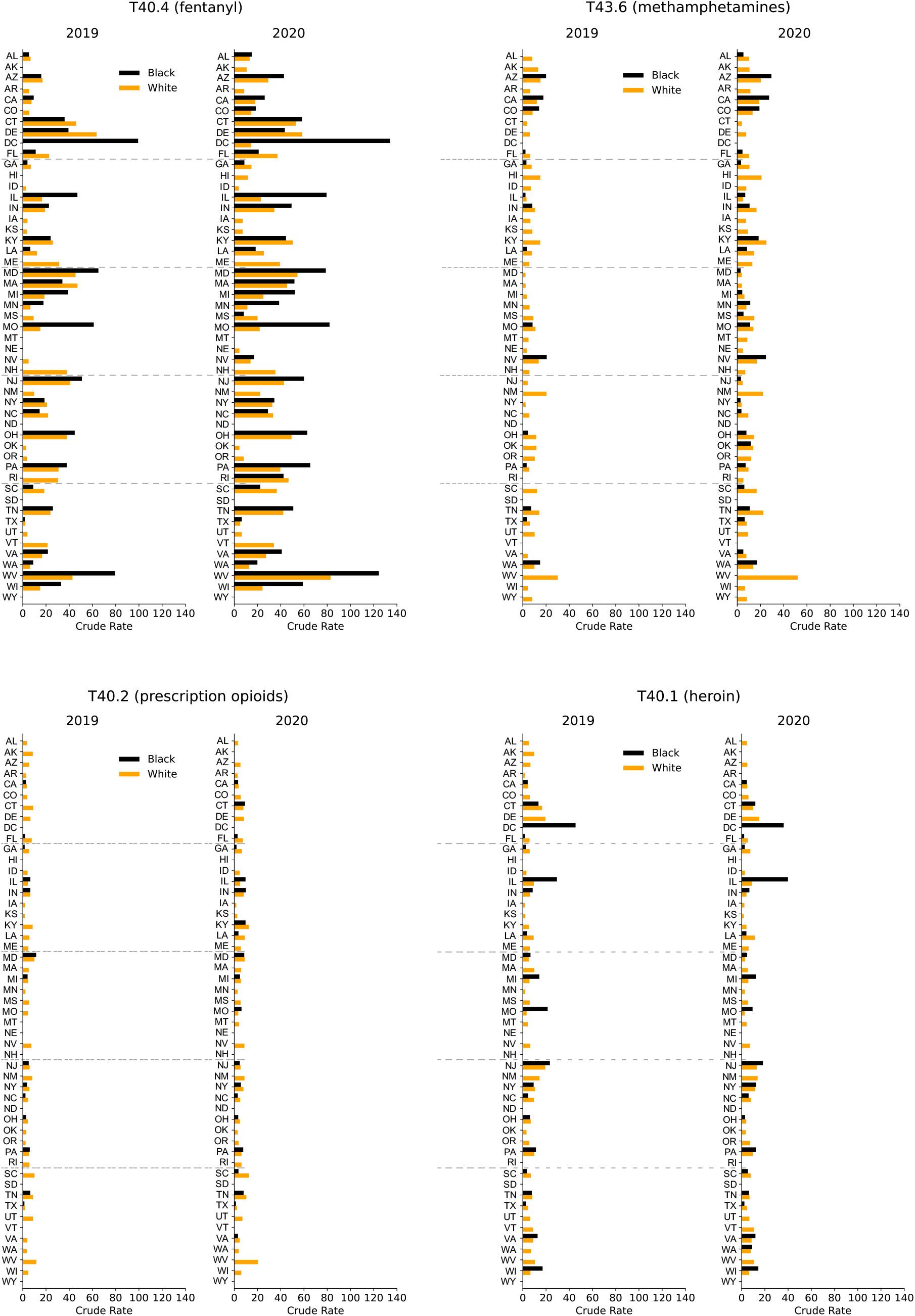
Crude rates for Black and White males across the United States associated with fentanyl (T40.4), methamphetamines (T43.6), prescription opioids (T40.2), heroin (T40.1) for the years 2019 and 2020. Note the racial imbalances and the increase in mortality in the District of Columbia for fentanyl and heroin.

Large racial disparities are observed in the District of Columbia for male heroin deaths as well. Although Black male mortality in the District of Columbia peaked at 55 cases per 100,000 in 2016, the 2020 male crude rate is still among the highest in the nation at 36. There is insufficient data for Whites. This pattern of elevated Black mortality and not statistically significant White overdoses is consistent over the entire 2013-2020 period. The state with the largest Black male crude rate in 2020 is Illinois, at 40, whereas the corresponding rate for Whites is 9, representing a 4.4 Black to White male ratio. Throughout 2013-2020 heroin crude rates in Illinois have been higher for Blacks than for Whites; while rates kept increasing or stabilized for Blacks, they began declining for Whites in 2017.

Other states with large racial gaps for fentanyl overdoses during 2020 are in the Midwest, and include Illinois, Minnesota, and Missouri, for both genders, consistent with 2019 data. The male racial gap increased in 2020 compared to 2019 in all states for which data is available except for West Virginia, Missouri and Kentucky, where it decreased slightly; for women the racial gap increased in most states, but decreased in ten mostly southern states including South Carolina, Louisiana and Florida. Much larger crude rates for Whites than Blacks are observed in 2020 for T43.6 and T40.2 in several southern states, including Georgia, South Carolina and Florida, consistent with 2019 data.

Finally, to study overall gender dynamics in each state, we combined all four drug categories and constructed the 2013-2020 time series. Overdose rates increased everywhere in 2020 compared to 2019, except for New Hampshire and South Dakota for both genders. For males, drug overdoses also decreased in Delaware, for females in Montana and New Jersey. The combined male-to-female overdose ratio increased in 36 states in 2020 compared to 2019. Significant upward shifts occurred in Kentucky, West Virginia and Colorado where the 2020 ratio was three times more the standard deviation derived from the 2013-2019 deaths. In all three states, male overdoses rose more than female overdoses. There were also 13 states where the male-to-female ratio decreased or remained comparable to 2019. The most significant downward shifts in the male-to-female ratios were recorded in Alaska (+16% males, +82% females) and Delaware (−3% males, +20% females), indicating relatively higher drug overdoses among females in these states.

## DISCUSSION

The year 2020 saw increases in drug-induced mortality across the United States compared to previous years. Mortalities for fentanyl and methamphetamines exceed projections from 2013-2019 (which already anticipated large increases) for both genders. Overall, 2013-2020 saw an explosion in overdose rates nationwide with male overdose by synthetic opioids increasing by 2,209%; and female rates by 991%. Methamphetamine rates follow similar patterns (+550% males, +486% females). Beginning in 2017, natural and semi-synthetic opioid overdose rates began decreasing for both genders, followed by a large, statistically significant resurgence in 2020; a similar pattern is observed for heroin for which the limited 2020 increase was not statistically significant. Male mortality exceeds female mortality in all states; the 2020 male-to-female overdose ratio nationwide was highest for heroin (3.1), followed by synthetic opioids (2.8), methamphetamines (2.6) and natural and semi-synthetic opioids (1.6). This is consistent with data from previous years and with the hypothesis that females consume (and die) more from natural and semi-synthetic opioids contained in prescription pain-killers which are easier and safer to acquire than street drugs [7].

A more nuanced picture emerges after racial stratifications are included. For example, crude rates for fentanyl overdoses among Black males and females have increased more than among Whites over the past few years; in 2018, and for the first time, Black male crude rates reached and later surpassed those of White males. Between 2013 and 2020 fentanyl crude rates for Black males rose nationwide by 6,980%; those for Black females rose by 2,140%. These increases are much larger than those recorded for the population at large (+2,209% male; +991% female) and for Whites (+ 2,033% male; +840% female). Similarly, the crude rate of heroin overdoses for the male and female population at large is decreasing within the United States and among Whites, however it is still increasing among Black males.

Combined gender, race and geographical stratifications are important as they may shift our perception of the most at risk groups [12, 13]. For example, in 2020 the highest crude rate for fentanyl deaths is in West Virginia at 84 cases per 100,000 persons. In the District of Columbia it is 68. However, as discussed above, a deeper analysis reveals that the fentanyl epidemic is much more severe for Black males in the District of Columbia with 134 cases per 100,000 Black males; the White male crude rate is 14, indicating significant disparity between the two races. The crude rate for Black females in the District of Columbia is 23; there is insufficient data for White females. This imbalance is also evident in West Virginia where the fentanyl male crude rate is 125 for Blacks and 83 for Whites. The overall crude rate in West Virginia, at 84, is comparable to that of White males because of the smaller percentage of Blacks residing in the state, which heavily weighs the crude rate towards that of Whites. Our results reinforce the importance of including racial stratifications as they provide a better gauge of the severity of the drug epidemic.

Fentanyl crude rates for Black males exceeded those of White males in many other states where there is sufficient 2020 data to allow for racial stratification. Large disparities are seen in Illinois, Minnesota and Missouri. The trend is reversed in most Southern states where male crude rates remain higher for Whites than Blacks. The highest 2020 crude rate for Black females is in the District of Columbia, at 43. Deaths among White females are too low to yield a significant statistic. Large racial disparities also emerge for male heroin overdoses in Illinois, Missouri and Michigan.

Another important finding is that the year 2020 saw male overdose crude rates for all drug types and genders increasing more than female crude rates almost everywhere except for Alaska, where crude rates rose by 16% for males but by a very large 82% for females. In Delaware male crude rates decreased by 3% but female mortality rose by 20%.

Why do some jurisdictions display such wide racial and gender disparities, while in others overdose rates are more uniform? One step further in our analysis, and an attempt to resolve this question, would be to quantify other factors such as poverty, inequality indices, income, unemployment, drug availability, access to prevention and treatment (such as naloxone) in various regions, and determine which of these correlate the most with the observed crude rates. Such an analysis could help provide a clearer, more nuanced picture of the drug epidemic and help prioritize interventions.

## LIMITATIONS

Our analysis is based on four drug categories selected from the CDC WONDER database (T40.4, T40,2, T40.1, T43.6). They are the ones with the largest crude rates, allowing us to follow temporal trends and stratify deaths as much as possible by race and gender. Other categories such as T40.3 (methadone) or T40.5 (cocaine) were not included in this study due to lower mortality rates and may display different dynamics. We did not consider Hispanic origin, nor did we consider other races such Asian or Pacific Islander (except in Figure 1), or American Indian or Alaska natives. While still much lower than the mortality rates of Blacks and Whites, both Asian and American Indian mortality rates have been increasing in recent years. We did not include any stratification by age. Since WONDER suppresses entries where the number of deaths is less than 10, data has not been available for some combinations of states, genders and races with extremely low fatalities (typically females). Finally, if multiple drugs are found in a person who overdosed, the death is tallied in all respective categories, resulting in multiple counts.

## PUBLIC HEALTH IMPLICATIONS

There are several public health implications stemming from our work. First, although identifying the underlying causes of increasing overdoses in 2020 is beyond the scope of this work, it is reasonable to assume that pandemic-related anxieties have been major factors in rising mortality [14--16], together with the greater availability of cheaper (and potentially contaminated) drugs, and the ease of ordering drugs online [17]. Isolation may have also made it easier to abuse drugs solo, with less access to emergency resources and partners in case of overdose. Since decreasing prices, drug contamination, and easier off-the-street procurement methods may persist in a post-pandemic world, overdose rates may remain high in the coming years and efforts should be devoted to monitoring online and mail-order drug transactions. Second, we find that stratifying by race and gender may lead to surprising differences between demographic groups. We find this to be true for overdoses occurring even prior to the pandemic. Thus, in order to more efficiently implement ef prevention and harm reduction initiatives one must first understand the internal dynamics and barriers facing diverse groups vis-à-vis drug addiction [18-20]. Given that Black males are often the most-at-risk category, and that their overdose rates are increasing dramatically, dedicated programs should be tailored to this specific demographic along the lines recommended in [21]. These include educational preventive initiatives, awareness of overdose risks associated to fentanyl, naloxone distribution, access to treatment programs and post-detox support.

Data and codes for plotting are available at this link: XXX

## Data Availability

Data and codes for plotting are available at this link: https://github.com/dorsogna/drugs

## ACKNOWLEDGMENTS

This work was supported by the Army Research Office through Grant No. W911NF-18-1-0345, the NIH through Grant No. R01HL146552 (T.C.), and the NSF through Grant No. DMS-1814090 (M.R.D.).

Note. The funder had no role in the design and conduct of the study; the collection, management, analysis, and interpretation of the data; the preparation or review of the article; or the decision to submit the article for publication.

## REFERENCES

[1] Ahmad F. B., Rossen L. M., Sutton P. Provisional drug overdose death counts. National Center for Health Statistics (2021).

[2] Friedman J., Beletsky L., Schriger D. L. Overdose-related cardiac arrests observed by emergency medical services during the US COVID-19 epidemic. JAMA Psychiatry 78 (2021) 562–564.

[3] United Nation Office on Drugs and Crime (UNODC) COVID-19 and the drug supply chain: from production and trafficking to use, Vienna (2020).

[4] Hedegaard H., Miniño, A. M., Spencer, M.R., Warner, M. Drug overdose deaths in the United States, 1999–2020 NCHS Data Brief No. 428, December 2021.

[5] Mattson C. L., Tanz L. J., Quinn K., Kariisa M., Patel P., Davis N.L. Trends and geographic patterns in drug and synthetic opioid overdose deaths -- United States, 2013–2019. MMWR Morbidity and Mortality Weekly Reports 70 (2021) 202–207.

[6] Ho J. Y. Cycles of gender convergence and divergence in drug overdose mortality, Population and Development Review 46 (2020) 443–470.

[7] Becker J. B., Mc Clellan M.L., Reed, B. G. Sex differences, gender and addiction, Journal of Neuroscience Research 95 (2017) 136–147.

[8] Larochelle M. R., Slavova S., Root E. D., Feaster D. J., Ward P. J., Selk S.C., Knott C., Villani J., Samet J. H. Disparities in opioid overdose death trends by race/ethnicity, 2018–2019, from the HEALing Communities Study. American Journal of Public Health 111 (2021) 1851–1854.

[9] O’Donnell J., Tanz L. J., Gladden M., Davis N.L., Bitting J. Trends in and characteristics of drug overdose deaths involving illicitly manifactured fentanyls -- United States, 2019–2020. MMWR Morbidity and Mortality Weekly Reports 70 (2021) 1740–1746.

[10] Friedman J., Hansen H., Evaluation of increases in drug overdose mortality rates in the US by race and ethnicity before and during the COVID-19 pandemic. JAMA Psychiatry 79 (2022) 379–381.

[11] Wide-ranging online data for epidemiologic research (WONDER). Atlanta, GA: CDC, National Center for Health Statistics (2020). Available at http://wonder.cdc.gov.

[12] Friedman J., Hansen, H. Far From a “White Problem”: Responding to the overdose crisis as a racial justice issue. American Journal of Public Health 112 (2022), S30–S32.

[13] Hulsey J. N. Toward improved addiction treatment quality and access for Black patients. American Journal of Public Health 112 (2022) S21–S23.

[14] Cartus A. R., Li Y., Macmadu A., Goedel W. C., Allen B., Cerda M., Marshall B. D. L. Forecasted and observed drug overdose deaths in the US during the COVID-19 pandemic in 2020, JAMA Network Open 5 (2022) e223418.

[15] Sher L. The impact of the COVID-19 pandemic on suicide rates. QJM: An International Journal of Medicine113 (2020) 707–712.

[16] Woolf S. H., Chapman D. A., Sabo R. T., Weinberger D. M., Hill L. Excess deaths from COVID-19 and other causes, JAMA 324 (2020) 510–513.

[17] Shelley L., Fentanyl, COVID-19, and Public Health, World Medical and Health Policy 12 (2020) 390–397.

[18] Brady K. T., Randall C. L., Gender differences in substance use disorders, Addictive Disorders 22 (1999) 241–252.

[19] Hansen H., Jordan A., Plough A., Alegria M., Cunningham C., Ostrovsky A. Lessons for the opioid crisis - Integrating social determinants of health into clinical care. American Journal of Public Health, 112(S2) (2022) S109–S111.

[20] Lopez A. M., Thomann M., Dhatt Z., Ferrera, J., Al-Nassir M., Ambrose M., Sullivan S. Understanding racial inequities in the implementation of harm reduction initiatives. American Journal of Public Health, 112 (2022) S173–S181.

[21] Blackwood C. A., Cadet J. L. COVID-19 Pandemic and fentanyl use disorder in African Americans. Frontiers in Neuroscience, 15 (2021).

